# Temporal detection and phylogenetic assessment of SARS-CoV-2 in municipal wastewater

**DOI:** 10.1101/2020.04.15.20066746

**Authors:** Artem Nemudryi, Anna Nemudraia, Tanner Wiegand, Kevin Surya, Murat Buyukyoruk, Karl K Vanderwood, Royce Wilkinson, Blake Wiedenheft

## Abstract

SARS-CoV-2 has recently been detected in feces, which indicates that wastewater may be used to monitor viral prevalence in the community. Here we use RT-qPCR to monitor wastewater for SARS-CoV-2 RNA over a 52-day time course. We show that changes in SARS-CoV-2 RNA concentrations correlate with local COVID-19 epidemiological data (R_2_=0.9), though detection in wastewater trails symptom onset dates by 5-8 days. We determine a near complete (98.5%) SARS-CoV-2 genome sequence from the wastewater and use phylogenic analysis to infer viral ancestry. Collectively, this work demonstrates how wastewater can be used as a proxy to monitor viral prevalence in the community and how genome sequencing can be used for high-resolution genotyping of the predominant strains circulating in a community.

---

In late December of 2019, authorities from the Peoples Republic of China (PRC) announced an epidemic of pneumonia (WHO, 2019). A novel coronavirus (Severe Acute Respiratory Syndrome coronavirus 2, SARS-CoV-2) was identified as the etiologic agent and the disease has been named “coronavirus disease 2019” (COVID-19). The virus spread rapidly, first to Thailand, Japan, Korea, and Europe, and now to over 188 countries across all continents except Antarctica. The global total of infected individuals now exceeds six million (https://coronavirus.jhu.edu/).

Public health professionals around the world are working to limit the spread of SARS-CoV-2, and “flatten the curve”, which requires a reduction in cases from one day to the next. However, SARS-CoV-2 containment has been outpaced by viral spread and limited resources for testing. Moreover, mounting evidence suggests that the virus is not only spread by aerosols but may also be transmitted via feces. Both viral RNA and infectious virus have been detected in the stool of COVID-19 patients (Hindson, 2020; Lodder and de Roda Husman, 2020; Tang et al., 2020a; Wang et al., 2020; Wu et al., 2020c; Xiao et al., 2020; Xu, 2020). This has important implications for the spread of the virus and suggests that wastewater may be used to monitor progression or abatement of viral spread at the community level (Mallapaty, 2020). To test this hypothesis, we collected samples of untreated wastewater from the municipal wastewater treatment plant in Bozeman, Montana (USA). Samples were collected on 12 different days over the course of a 52-day period, using two different collection methods. The samples were filtered and concentrated prior to RNA extraction. Isolated RNA was used as a template for one-step reverse transcription quantitative polymerase chain reaction (RT-qPCR), performed according to CDC guidelines (https://www.fda.gov/media/134922/download). Each RT-qPCR reaction was performed using two primer pairs (i.e., N1, and N2), which target distinct regions of the nucleocapsid (N) gene from SARS-CoV-2 (**Fig 1 and Supplemental Fig S1**). The first two influent samples were collected manually at the headworks, using a sampling stick on the mornings of March 23^rd^ and 27^th^, respectively. Both samples tested positive for SARS-CoV-2 RNA (**Fig 1A**). To minimize inherent variability that comes with collecting a single sample from a large heterogeneous volume at any given point in time, subsequent sampling was performed using an autosampler that collects a volume proportional to flow for 24-hours. This composite sample reflects average characteristics of the wastewater over the previous day. Similar to the first two timepoints, composite samples collected in late March and early April tested positive for SARS-CoV-2, though concentrations of viral RNA steadily declined and then dropped below the limit of detection (**Fig 1A and Supplemental Table S1**). To better understand how SARS-CoV-2 RNA concentrations measured in the wastewater compare to individual SARS-CoV-2 infections in the community, we gathered symptom onset data from patients that tested positive for SARS-CoV-2 in Bozeman. This comparison was performed using a cross-correlation analysis, which measures similarity as a function of displacing one dataset relative to the other. We compared concentration of SARS-CoV-2 RNA determined using the N1 or N2 primer pairs, to symptom onset data that was collected by a retrospective survey of individuals that tested positive for SARS-CoV-2. N1 has a maximum positive correlation (0.736) with symptom onset when we account for a of 5-day lag, while N2 has a maximum positive correlation (0.589) with symptom onset with a 7-day lag (**Fig 1B**). We hypothesized that the number of COVID-19 cases should have a linear relationship with viral RNA copies in wastewater. To test this hypothesis, we performed a regression analysis. In agreement with the cross-correlation analysis, linear regression reveals that changes in SARS-CoV-2 RNA concentrations are best predicted by symptom onset dates with a 5-to 8-day lag (**Fig 1C**). However, an 8-day lag for N1 (R_2_ = 0.93) and a 7-day lag for N2 (R_2_ = 0.9) result in the best fits for the linear models (**Fig 1D, E**). This observation agrees with clinical data showing a delay between symptom onset, and detection of SARS-CoV-2 in respiratory and stool samples of COVID-19 patients (Cheung et al., 2020; Wu et al., 2020c).

**Fig 1.**
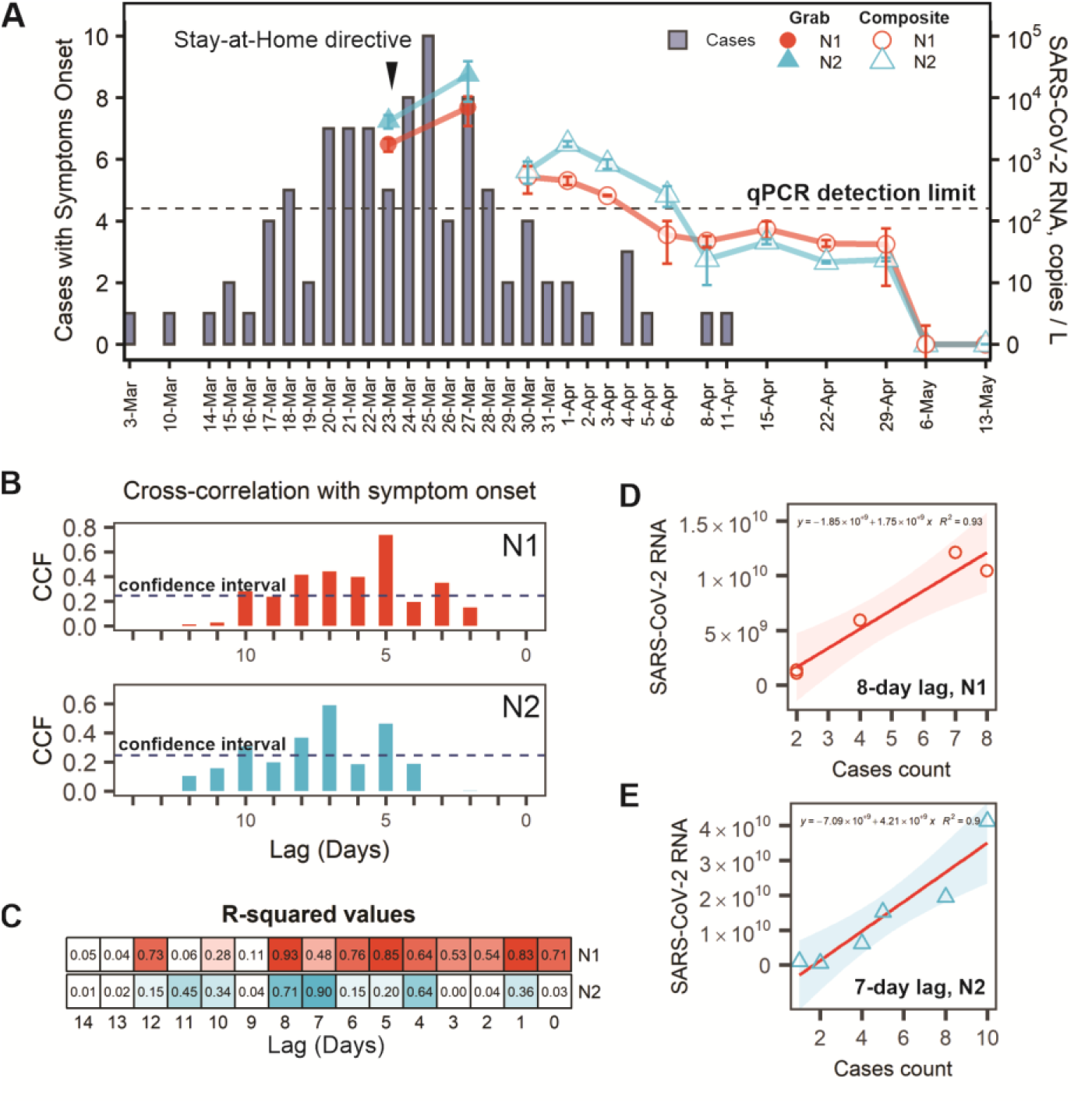
Detection and quantification of SARS-CoV-2 in wastewater and in the community. **A)** SARS-CoV-2 RT-qPCR tests were performed according to CDC guidelines and protocols. Detection included two primer pairs, each targeting distinct regions of the nucleocapsid (N) gene from SARS-CoV-2 (i.e., N1, and N2). Three 0.5-liter samples were collected manually (grab) or subsampled from a 24-hour composite. SARS-CoV-2 RNA concentrations (copies per liter) were estimated using a standard curve. The limit of detection in this assay is 10 copies per 20 µl qPCR reaction. For composite samples, concentrations were normalized according to the total daily volume (see Methods). Temporal dynamics of SARS-CoV-2 RNA (line graph) superimposed on the epidemiological data (bar plot). Symptom onset data was collected by interviewing COVID-19 patients. Bars represent number of patients who reported symptom onset on the specified day. **B)** Cross-correlation of epidemiological data and SARS-CoV-2 RNA levels in wastewater. Values of cross-correlation function (CCF) are offset by 0-to 14-days. The 95% confidence interval is shown with dashed blue line. CCFs above this line are statistically significant. **C)** The relationship between symptom onset and SARS-CoV-2 RNA measured in wastewater was modeled using linear regression. Goodness-of-fit (R-squared values) metrics are shown for displacing RT-qPCR data relative to symptom onset by 0-to 14-days. **D and E)** Linear regression between symptom onset and total SARS-CoV-2 RNA in wastewater adjusted for 8-day and 7-day lags for N1 and N2, respectively. Pink and blue shadows define the 95% confidence interval.

To verify that the RT-qPCR results reflect bona fide detection of SARS-CoV-2 rather than priming from an unintended template, we repeated the PCR using 10 primer pairs that tile across the SARS-CoV-2 genome (Quick, 2020). These primers were designed to target conserved regions of the genome that flank polymorphic sites that have been used to trace viral ancestry and geographic origins (Bedford et al., 2020; Quick, 2020) (**Supplemental Fig S2A**). RNA isolated from the Bozeman waste stream on March 27^th^ was used as a template for these RT-PCR reactions, and all 10 primer pairs produced PCR products of the expected sizes (**Supplemental Table S2 and Fig S2B**). PCR products were sequenced using Sanger methods and the reads were aligned to the reference genome using MUSCLE (Edgar, 2004; Wu et al., 2020b). We observed no sequence heterogeneity in redundant reads derived from each location of the genome (**Supplemental Fig S2C**), which suggests the predominance of a single SARS-CoV-2 genotype in the waste stream at the time of sampling. We then used this same RNA sample to determine a near complete (98.5%) SARS-CoV-2 genome sequence using a long read sequencing platform (Quick, 2020).

Efforts to understand the origins and evolution of SARS-CoV-2 have resulted in ∼35,000 genome sequences from 86 countries as of May 30^th^, 2020 (https://www.gisaid.org/). Phylogenetic analyses of these sequences have enabled molecular tracking of viral spread (Castillo et al., 2020; Fauver et al., 2020; Gonzalez-Reiche et al., 2020; Stefanelli et al., 2020; van Dorp et al., 2020; Zehender et al., 2020). To determine the ancestry of the predominant SARS-CoV-2 strain circulating in Bozeman’s wastewater on March 27th, we determined the genome sequence using Oxford Nanopore.

Sequencing resulted in ∼700,000 reads. Quality control and base calling were performed with MinKNOW v19.06.8 in High Accuracy mode (Oxford Nanopore Technologies) and the sequences were assembled using the bioinformatic pipeline from ARTIC Network (https://artic.network/ncov-2019). This approach resulted in a single viral contig with 6,875X average sequencing depth that covered 98.5% of the SARS-CoV-2 reference genome. Unsequenced regions of the genome include the 5’- and 3’-ends, and a stretch of 170 bases (22,346 – 22,515 in the reference genome), that likely had too few reads for basecalling due to PCR bias (Itokawa et al., 2020). This genome was aligned to 14,970 SARS-CoV-2 genomes from 74 different countries (Global Initiative on Sharing All Influenza Data; https://www.gisaid.org/). The resulting alignment was used to build a phylogenetic tree (**Fig 2A)**, which indicates that the predominant SARS-CoV-2 genotype in Bozeman’s wastewater is most closely related to genomes from California (USA) and Victoria (Australia) (**Fig 2B**). In total, 11 mutations distinguish the Bozeman wastewater SARS-CoV-2 sequence from the Wuhan-Hu-1/2019 reference sequence, 10 of these 11 mutations are also present in sequences from California and nine of these 11 mutations are also in an isolate from Victoria (**Fig 2B and Supplemental Fig S3A**). To determine how these sequence variations may have accumulated over space and time, we mapped each mutation onto the phylogenetic tree of SARS-CoV-2 sequences (**Supplemental Fig 3B**). Mutations that do not confer a fitness defect are preserved in viral progeny, and thus serve as genetic landmarks that can be used to trace viral ancestry (**Supplemental Fig 3B**). This analysis indicates that the predominate viral stain circulating in Bozeman is most closely related to a strain circulating in California and that the Bozeman isolate acquired an additional mutation (A23122T) that does not co-occur with the other ten mutations. Thus, mutation A23122T may provide a unique genetic signature that can be linked to Bozeman. While this sequencing approach reveals the genetic history of the dominant SARS-CoV-2 variant circulating in Bozeman wastewater at the time of sampling, it does not measure the fitness of this or any of the mutations associated with distinct geographic locations. We anticipate that temporal genome sequencing will help identify dominant strains of the virus circulating in a specific community over time.

**Fig 2.**
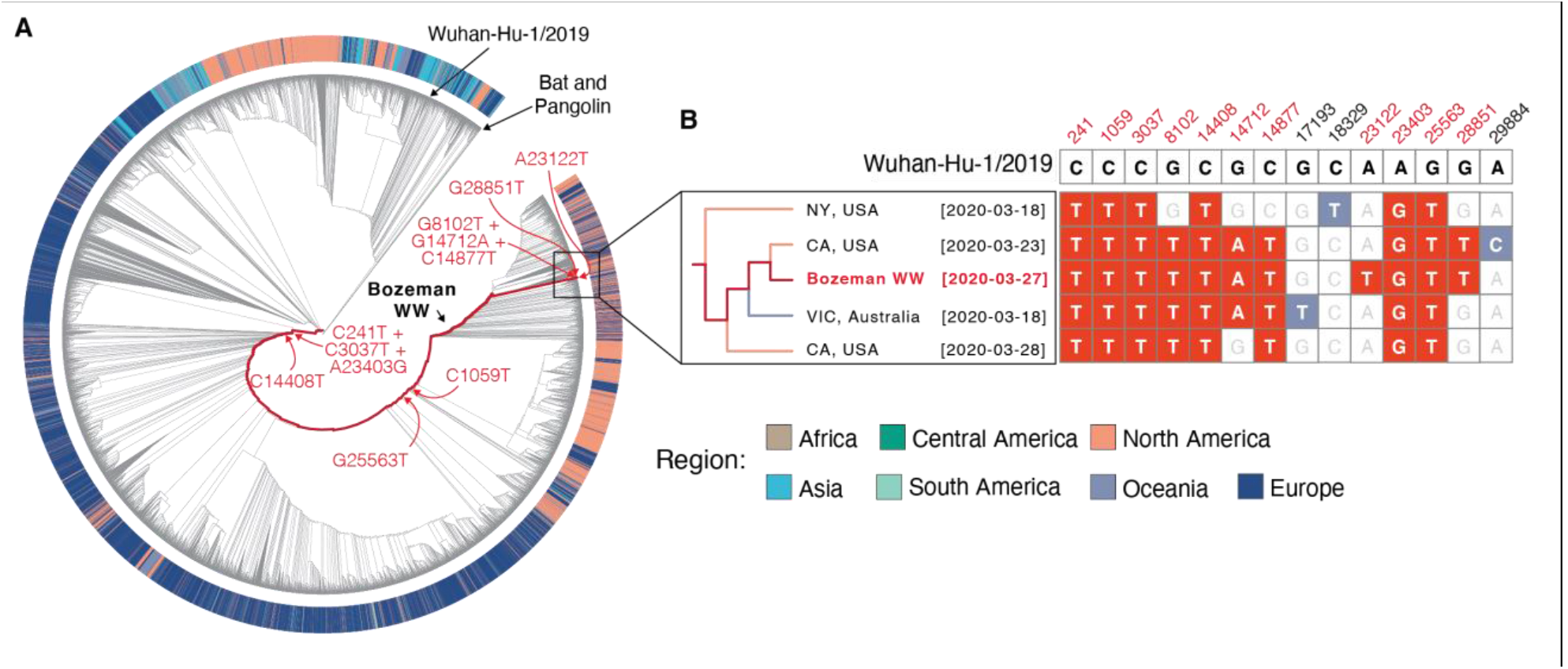
Phylogenetic analysis of SARS-CoV-2 sequences from wastewater. **A)** Maximum-likelihood phylogeny of the SARS-CoV-2 related lineage (n = 14,971 sequences). Phylogenetic history of SARS-CoV-2 strain sequenced from Bozeman’s wastewater is shown in crimson. Outer ring colored according to regions of the world where the samples were isolated. Tree is rooted relative to RaTG13 genome (a bat coronavirus with 96% sequence similarity to SARS-CoV-2; Genbank: MN996532.1). Mutations that occurred over space and time are shown in red. **B)** Inset from panel B: sequences isolated from Bozeman wastewater clade with sequences of American and Australian origin (left). Sequences are named according to geographic origin and viral isolation date. Comparison of mutations in sequences shown in inset (right). The Wuhan reference sequence for each of the positions where mutations occur is shown across the top. Mutated positions and bases present in Bozeman wastewater (WW) sequence are shown in red, bases matching Wuhan reference sequence are shown in white, and mutations not present in Bozeman WW sequence are shown in blue.

Collectively, the results presented here demonstrate that wastewater monitoring for SARS-CoV-2 RNA by RT-qPCR correlates with prevalence of viral infections in the community (**Fig 1**). Our data indicate that symptom onset precedes detection of SARS-CoV-2 RNA in wastewater by 5-8 days. However, patient testing or hospital admission typically occurs 3-9 days after symptoms onset (Garg et al., 2020). Thus, an increase of SARS-CoV-2 RNA in wastewater may coincide or even precede by a day or two the detection of new COVID-19 cases in a community. Furthermore, wastewater surveillance may capture mild and asymptomatic infections that are actively shedding virus, which could be used to and alert Public Health officials about emerging undetected transmission events (Tang et al., 2020b).

While this paper was under review, several preprints and publications have reported similar findings from wastewater system around the world (Ahmed et al., 2020; La Rosa et al., 2020; Medema et al., 2020; Randazzo et al., 2020; Rimoldi et al., 2020; Wu et al., 2020a; Wurtzer et al., 2020). Despite SARS-CoV-2 RNA being present in the wastewater, and viral RNA and infectious virus found feces, infectious virus has not been isolated directly from wastewater (Rimoldi et al., 2020). The study presented here complements the rapidly emerging body of work by providing an important link between sewage surveillance, COVID-19 epidemiology and tracing SARS-CoV-2 spread patterns with genome sequencing.

## Data Availability

Data are is available on request from the authors. SARS-CoV-2 genome sequence from wastewater sequence was uploaded to GISAID (https://www.gisaid.org/), accession ID: EPI_ISL_437434.

## ACKNOWLEDGEMENTS

Research in the Wiedenheft lab is supported by the National Institutes of Health (1R35GM134867), the Montana State University Agricultural Experimental Station, the MJ Murdock Charitable Trust, the Gianforte Foundation and the MSU Office of the Vice President for Research. We are grateful to Josh French, Justin Roberts, and the other dedicated wastewater technicians that made this work possible. We gratefully acknowledge the authors, originating and submitting laboratories of the sequences from GISAID’s EpiFlu™ Database (**Supplemental Table S3**).

## METHODS

### Wastewater sampling

Wastewater samples were collected at the Bozeman Water Reclamation Facility (BWRF) that receives and treats domestic, commercial, and industrial wastewater from the City of Bozeman, Montana (USA). Wastewater is sourced from the city limits (∼60 km^2^, 49’831 population) with an average flow rate of ∼2.31 × 10^4^ m^3^ / d. Grab samples were collected in triplicates with 15 second intervals from the influent, immediately downstream of the 6-mm screen and grit washer, which remove large solids and heavy inorganic particles. Composite samples were collected from raw influent with automatic flow proportional sampler Liquistation CSF34 (Endress+Hauser) located at the entrance to the facility downstream of a rock trap. Autosampler was set to collect 150 mL of influent per 150’000 gal of flow (∼ 5.68 × 10^5^ L) 7 AM to 7 AM. During collection temperature was kept +2 to +6°C, and samples were stored at +4°C before processing (2-3 h). The composite sample was subsampled in three 500 mL aliquots. Samples were collected three times a week during the outbreak, and once a week after April 8^th^.

### Wastewater sample processing and RNA extraction

Each wastewater sample (500 mL) was sequentially filtered through 20 µM, 5 µM (Sartorius Biolab Products) and 0.45 µM (Pall Corporation) membrane filters and concentrated down to 150-200 µL using Corning Spin-X UF concentrators with 100 kDa molecular weight cut-off. Total RNA from concentrated samples was extracted with RNeasy Mini Kit (Qiagen) and eluted with 40 µL of RNase free buffer. This RNA was used as a template for RT-qPCR.

### Reverse Transcription quantitative PCR (RT-qPCR)

RT-qPCR was performed using two primers pairs (N1 and N2) and probes from 2019-nCoV CDC EUA Kit (IDT#10006606). SARS-CoV-2 in wastewater was detected and quantified using one-step RT-qPCR in ABI 7500 Fast Real-Time PCR System according to CDC guidelines and protocols (https://www.fda.gov/media/134922/download). 20 µL reactions included 8.5 µL of Nuclease-free Water, 1.5 µL of Primer and Probe mix, 5 µL of TaqPath 1-Step RT-qPCR Master Mix (ThermoFisher, A15299) and 5 µL of the template. Nuclease-free water was used as negative template control (NTC).

Amplification was performed as follows: 25°C for 2 min, 50°C for 15 min, 95 °C for 2 min followed by 45 cycles of 95 °C for 3 s and 55 °C for 30 s. To quantify viral genome copy numbers in the samples, standard curves for N1 and N2 were generated using a dilution series of a positive template control (PTC) plasmid (IDT#10006625) with concentrations ranging from 10 to 10,000 copies per reaction. Three technical replicates were performed at each dilution. The limit of detection was 10 copies of the control plasmid. The NTC showed no amplification over the 40 cycles of qPCR.

Run data was analyzed in SDS software v1.4 (Applied Biosystems). Threshold cycle (Ct) values were determined by manually adjusting the threshold to fall within exponential phase of the fluorescence curves and above any background signal. Ct values of PTC dilutions were plotted against log_10_(copy number) to generate standard curves. Linear regression analysis was performed in RStudio v1.2.1335 and the trend line equation (Ct = [*slope*] × [log_10_(copy number)] + b) was used to calculate copy numbers from mean Ct values of technical replicates for each biological replicate. Primer efficiencies calculated as E = (10^(−1/[*slope*])^ – 1) × 100% were 150.36 ± 12.13% for N1 and 129.45 ± 25.5% for N2 (n = 7 runs, mean ± sd).

### RT-PCR and SARS-CoV-2 genome sequencing

Reverse transcription was performed with 10 µL of RNA from SARS-CoV-2 positive wastewater sample using SuperScript™ III Reverse Transcriptase (Thermo Fisher Scientific) according to the supplier’s protocol.

The amplicon library for SARS-CoV-2 whole genome sequencing on Oxford Nanopore was generated as described in protocol developed by ARTIC Network (https://artic.network/ncov-2019) (Quick, 2020). Briefly, V3 primer pools containing 110 and 108 primers were used for the multiplex PCR (https://artic.network/ncov-2019). PCR reactions were performed using Q5 High-Fidelity DNA Polymerase (New England Biolabs) with the following thermocycling conditions: 98°C for 2min, 35 cycles of 98°C for 15 s and 65°C for 5 min, 35 cycles. Two resulting amplicon pools were combined and used for library preparation pipeline that included end preparation and Nanopore adaptors ligation. 20 ng of final library DNA was loaded onto the MinION flowcell for sequencing. A total of 304.77 Mb of raw sequencing data was collected.

PCR products used for Sanger sequencing were generated with a subset of primers from ARTIC V3 pools (**Supplemental Table S2**). PCR reactions were performed as described above. PCR products were analyzed on 1% agarose gels stained with SYBR Safe (Thermo Fisher Scientific), remaining DNA was purified using DNA Clean & Concentrator™ kit (Zymo Research) and sent to Psomagen for Sanger sequencing. Each PCR product was sequenced with both forward and reverse primers used for PCR.

### SARS-CoV-2 genome assembly

Nanopore raw reads (304.77 Mb) were basecalled with MinKNOW software in high-accuracy mode. Successfully basecalled reads (273.8 Mb) were further analyzed using the ARTIC bioinformatic pipeline for COVID-19 (https://artic.network/ncov-2019). Consensus sequence was generated with minimap2 and single nucleotide variants were called with nanopolish (both integrated in the pipeline) relative to Wuhan-Hu-1/2019 reference genome (Li, 2018; Quick et al., 2016; Wu et al., 2020b). The resulting assembly had nearly complete genome coverage (98.51%) with 6,875X average sequencing depth. Regions of the genome that were not captured by this sequencing method include 5’ and 3’ ends of the genome and a stretch of 170 nucleotides (22,346 – 22,515 nucleotide positions in reference genome), presumably due to amplicon drop-out. Consensus sequence was upload to GISAID (https://www.gisaid.org/), accession ID: EPI_ISL_437434.

### Phylogenetic and Position-specific Mutation Analysis

Phylogenetic analysis was performed by aligning the consensus sequence to 14,970 SARS-CoV-2 genomes retrieved from GISAID on 5/5/2020, 8:25:22 AM (https://www.gisaid.org/), using the FFT-NS-2 setting in MAFFT v7.429 (Katoh et al., 2019; Shu and McCauley, 2017). Columns composed of more than 70% gaps were removed with trimAl v1.2rev59 (Capella-Gutierrez et al., 2009).

A maximum-likelihood phylogenetic tree was constructed from this alignment using IQTree in the Augur utility of Nextstrain (Hadfield et al., 2018; Minh et al., 2020). The APE v5.3 package in R was used to re-root the tree relative to RaTG13 bat coronavirus genome sequence (Paradis and Schliep, 2019), and the tree was plotted using ggtree v3.10 package in R (Yu et al., 2017). The subtree, visualized in Figure 2B, was rendered in FigTree v1.4.4 (Rambaut, 2017).

Position specific mutation analysis was conducted in R using the BioStrings package (Pagès et al., 2019), and chromatograms of Sanger sequencing reads were rendered in SnapGene (GSL Biotech; available at snapgene.com).

### Symptoms onset and contact tracing data collection

Suspect cases of COVID-19 were tested in a CLIA lab and instructed to self-quarantine until notified of the RT-qPCR test results. All laboratory confirmed positive cases of COVID-19 were contacted via telephone by local public health nurses to complete contact tracing. During this interview, the nurses collected recorded symptoms, symptom onset date, travel history, contact with other known laboratory confirmed cases, close contacts and activities on the two days before symptom onset up until notification of a positive test. Data collection was conducted as part of a public health response. The study was reviewed by the Montana State University Institutional Review Board (IRB) For the Protection of Human Subjects (FWA 00000165) and was exempt from IRB oversight in accordance with Code of Federal regulations, Part 46, section 101. All necessary patient/participant consent has been obtained and the appropriate institutional forms have been archived.

### Quantification and Statistical Analyses

All statistical analyses were performed in RStudio v1.2.1335. Data in figures are shown as mean of three biological replicates (each with two technical replicates) ± standard error of mean (sem). Estimated copy numbers in RT-qPCR reactions were used to calculate titers per liter of wastewater for each biological replicate. Viral RNA concentrations in the composite samples were normalized ([SARS-CoV-2 concentration]_Normalized_ = [SARS-CoV-2 concentration] × (Daily flow / Average flow)). Cross correlation analysis was performed in RStudio using ccf function from stats R package. Linear regression analysis was performed by shifting the SARS-CoV-2 viral RNA data along the axis (**see Fig 1**) in one day increments and plotting against symptom onset. Data was plotted and analyzed using geom_smooth function from ggplot2 R package, with method = ‘lm’.

## Supplementary Data

**Supplementary Figure S1.**
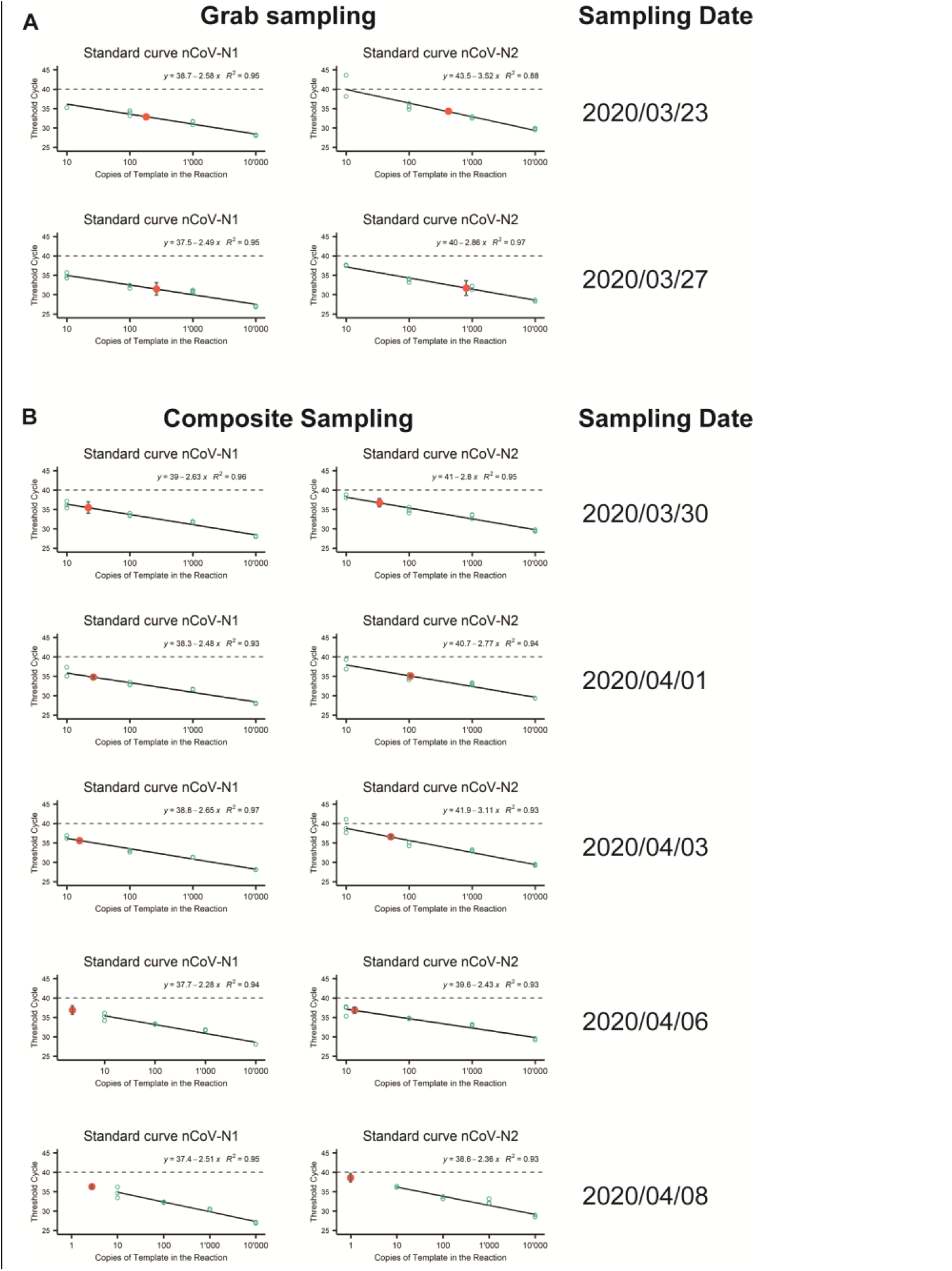
Standard curves for absolute quantification of SARS-CoV-2 titers. Standard curves were generated for each RT-qPCR run. A 10-fold dilution series of positive control plasmid (IDT#10006625) encoding for SARS-CoV-2 *N* gene were used, three technical replicates at each dilution (green circles). Data was plotted as Cycle Threshold (Ct) on y-axis versus log_10_(copy number) on x-axis. Trend lines were fitted to the data by linear regression analysis in RStudio v1.2.1335, Linear equations and R^2^ values are shown for each standard curve. Red dots show mean Ct values for wastewater samples. Error bars represent standard deviation of Ct values.

**Supplementary Figure S2.**
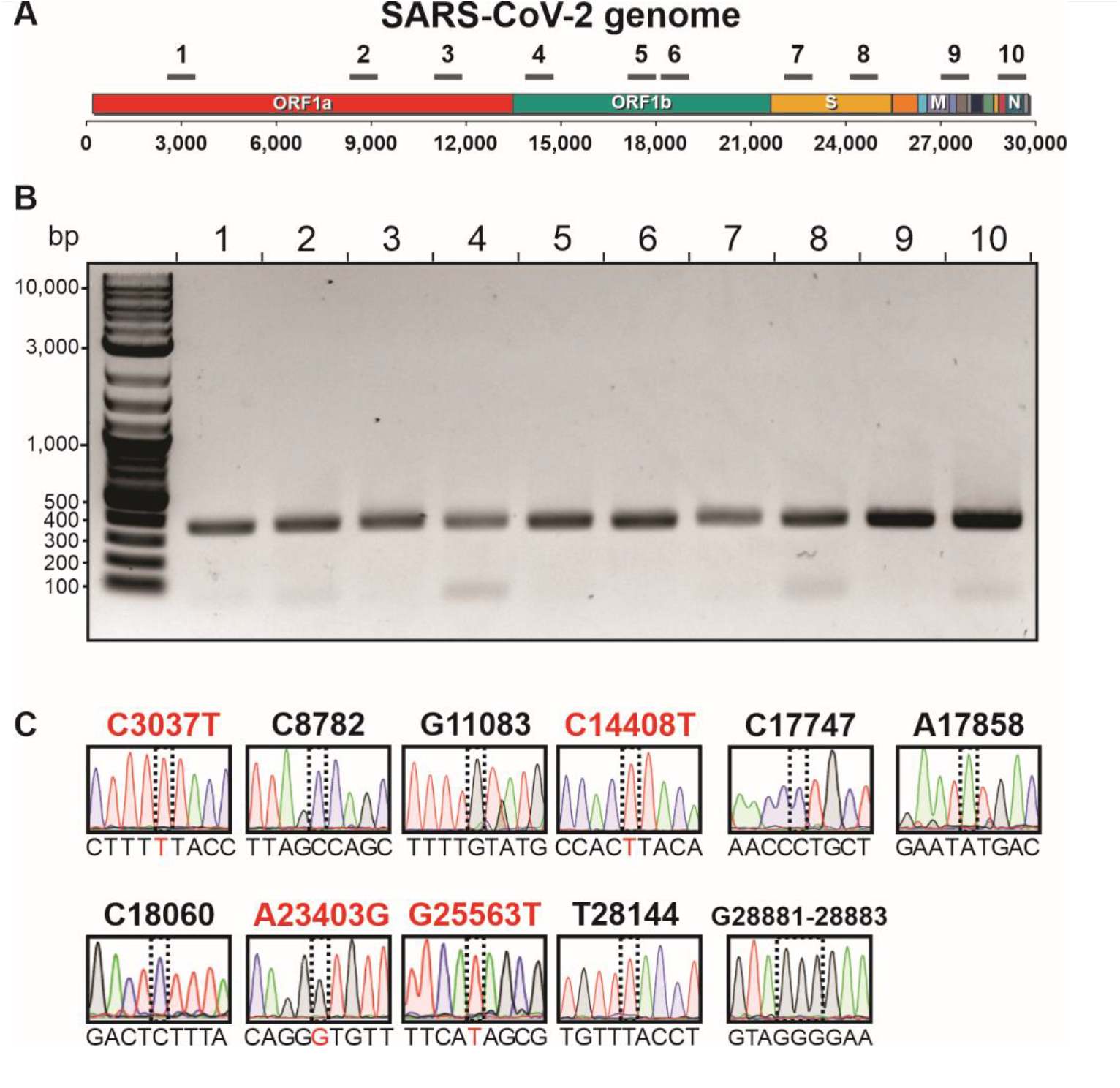
**A)** Map of the SARS-CoV-2 genome. The solid black lines indicate the approximate location of 10 amplicons spanning phylogenetically informative regions of the genome. **B)** Agarose gel of the corresponding PCR products. **C)** Ten PCR products shown in **B** were gel-purified and used to sequence 13 polymorphic sites in SARS-CoV-2 genome (Hadfield et al., 2018; Tang et al., 2020a). Sanger traces (9-bp windows) are shown for each polymorphic site (dotted boxes). Nucleotide numbering is based on the reference genome Wuhan-Hu-1/2019 (Wu et al., 2020b). Highlighted in red are nucleotide substitutions relative to the reference.

**Supplementary Figure S3.**
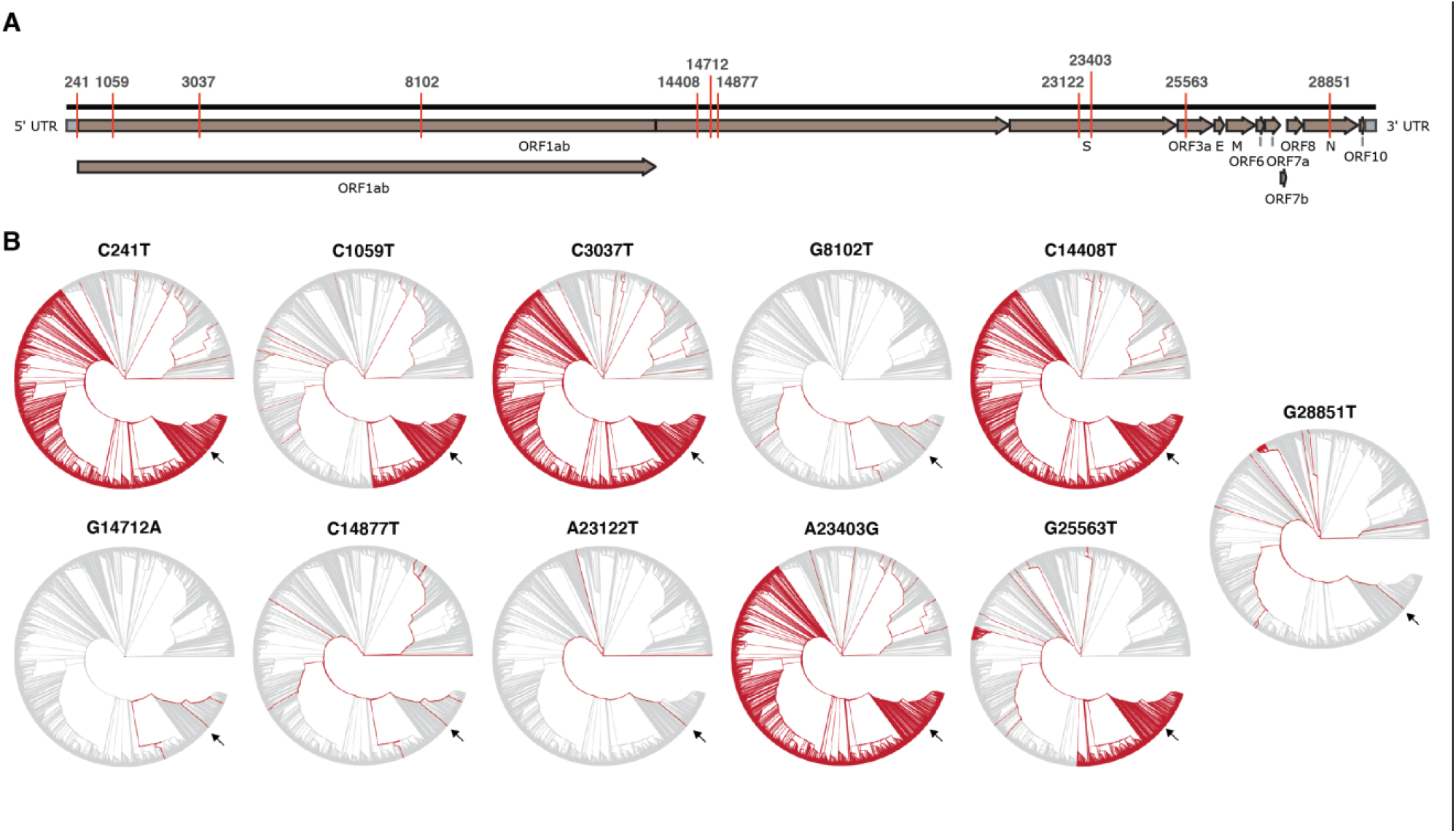
Mutations in Bozeman WW Sequence. **A)** Genetic map of SARS-CoV-2 with mutations present in Bozeman wastewater sequence highlighted. **B)** Phylogenetic tree with each mutation present in Bozeman wastewater mapped in red. The SARS-CoV-2 sequence from Bozeman is indicated by black arrow.

**Supplemental Table S1.**
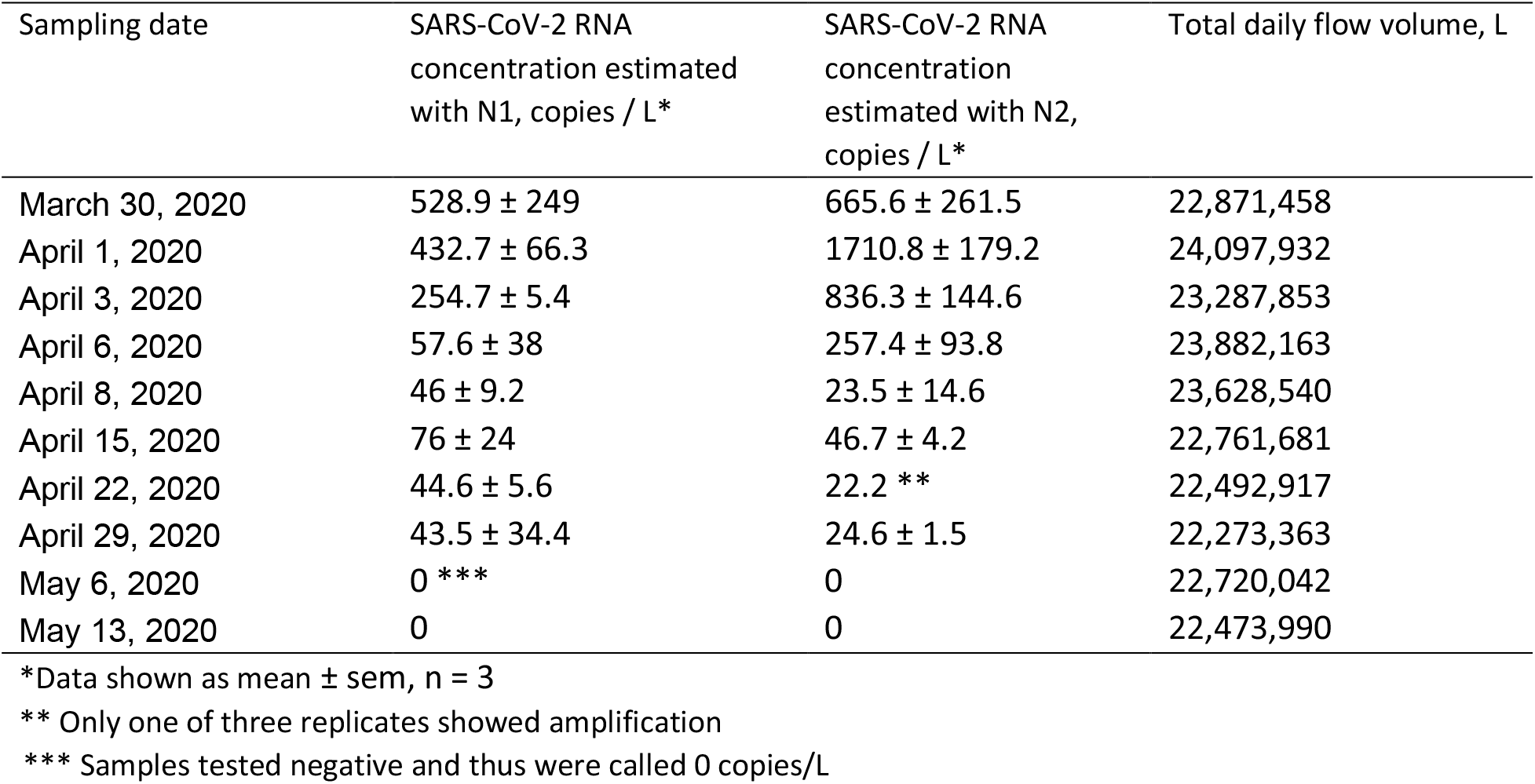
Total wastewater flow volumes and SARS-CoV-2 RNA concentration in composite samples.

| Sampling date | SARS-CoV-2 RNA concentration estimated with N1, copies / L * | SARS-CoV-2 RNA concentration estimated with N2, copies / L * | Total daily flow volume, L |
| --- | --- | --- | --- |
| March 30, 2020 | 528.9 ± 249 | 665.6 ± 261.5 | 22,871,458 |
| April 1, 2020 | 432.7 ± 66.3 | 1710.8 ± 179.2 | 24,097,932 |
| April 3, 2020 | 254.7 ± 5.4 | 836.3 ± 144.6 | 23,287,853 |
| April 6, 2020 | 57.6 ± 38 | 257.4 ± 93.8 | 23,882,163 |
| April 8, 2020 | 46 ± 9.2 | 23.5 ± 14.6 | 23,628,540 |
| April 15, 2020 | 76 ± 24 | 46.7 ± 4.2 | 22,761,681 |
| April 22, 2020 | 44.6 ± 5.6 | 22.2 ** | 22,492,917 |
| April 29, 2020 | 43.5 ± 34.4 | 24.6 ± 1.5 | 22,273,363 |
| May 6, 2020 | 0 *** | 0 | 22,720,042 |
| May 13, 2020 | 0 | 0 | 22,473,990 |

\*Data shown as mean ± sem, n = 3
\*\* Only one of three replicates showed amplification
\*\*\* Samples tested negative and thus were called 0 copies/L

**Supplemental Table S2.**
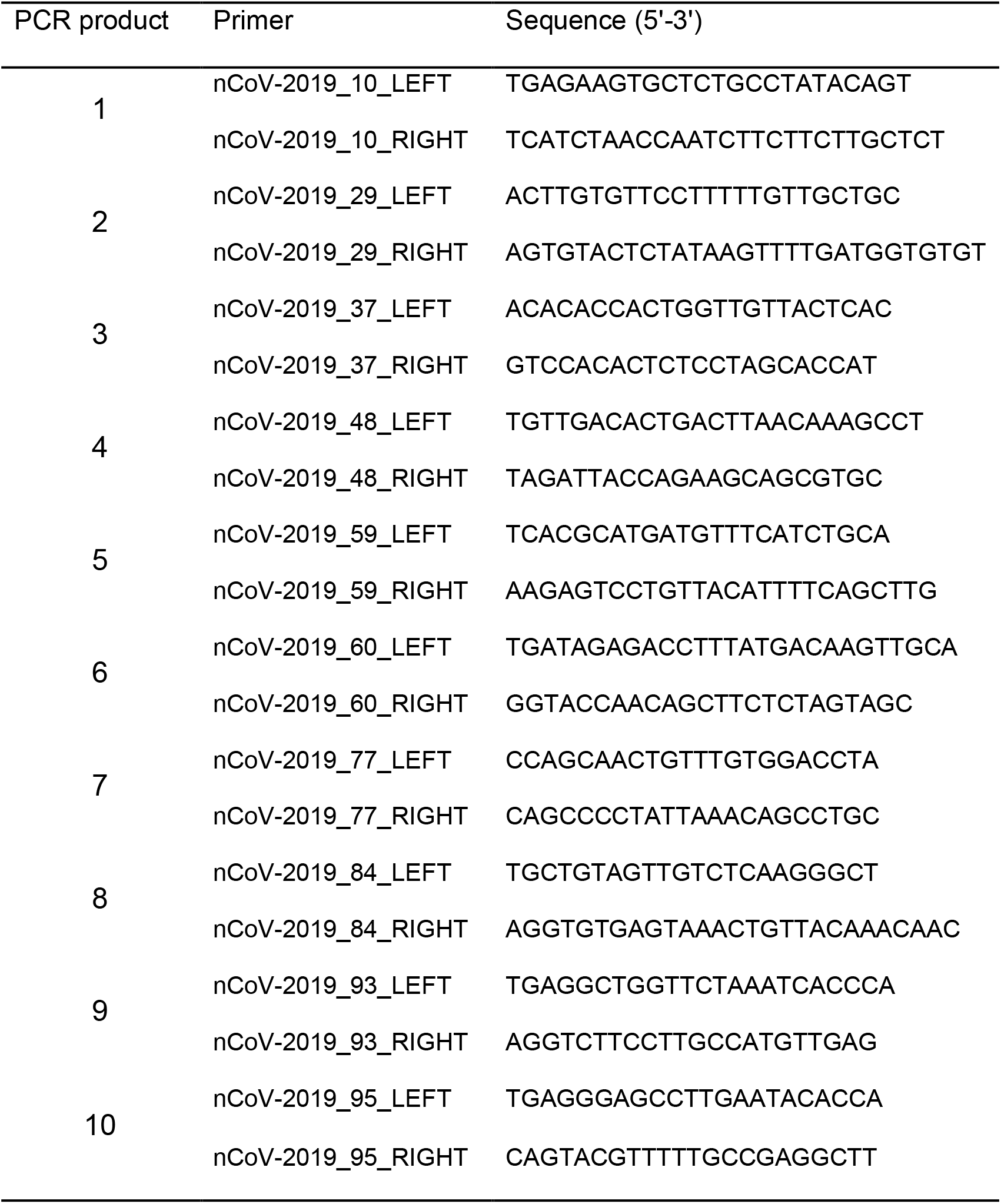
Primer design

